# Hydroxychloroquine serum concentrations in non-critical care patients infected with SARS-CoV-2

**DOI:** 10.1101/2020.06.23.20137992

**Authors:** Alasdair MacGowan, Fergus Hamilton, Mark Bayliss, Liam Read, Marie Attwood, Alan Noel, Sally Grier, Anna Morley, David Arnold, Nicholas Maskell

## Abstract

Hydroxychloroquine(HCQ) has been widely used to treat SARS-CoV-2 infection however HCQ pharmacokinetics in this condition have not been studied in non-critical care patient groups. Here we report the serum concentrations of HCQ in a small cohort of patients treated with HCQ as part of the RECOVERY trial.

## Introduction

Hydroxychloroquine (HCQ) has been widely used to treat infection with SARS-CoV-2 but most recently has been shown to be of no benefit in a large randomised clinical trial (RECOVERY 2020). Detailed information on the pharmacodynamics of HCQ is lacking but measures of the *in vitro* activity of HCQ against SARS-CoV-2 are available as well as pharmacokinetic data derived from analysis of patients with conditions other than SARS-CoV-2 infection who received HCQ. HCQ is much less active against SARS-CoV-2 *in vitro* than it is against Plasmodium species. Pharmacokinetic data from patients with SARS-CoV-2 is sparse and absent for those outside critical care (Perinel et al, 2020). Here we report data on the serum concentrations of HCQ in patients who were recruited into the Randomised Evaluation of COVID-19 Therapy (RECOVERY) Trial. Permission to take and use stored serum was ethically approved as part of the Diagnostic and Severity markers in COVID-19 to Enable Rapid triage (DISCOVER) Study. Patients received HCQ at a dose of 800mg 6 hrly apart, then 400mg 6 hours later followed by 400mg 12 hrly subsequently.

## Methods and Results

HCQ was assayed using a recently developed LC-MS/MS method. Serum was deproteinised with acetonitrile after addition of the internal standard, chloroquine. The method was validated over a working range of 0.005 to 1 mg/L. There was no evidence of drug loss in gel separated tubes compared to plain tubes or after 24 hr at room temperature. The mean blood/plasma partitioning ratio was 1.64 ± 0.25.

Seven patients were recruited into the HCQ treatment arm. Subsequently one patient (male, aged 58 years) had no blood samples drawn and a second (male, aged 84 years) only received a single dose of 800mg. Of the five remaining, two were male, three female with a mean ± std (min-max) age of 57.4 ± 20.7 (26-83) and weight of 70.6 ± 4.2 (65.4-75.0) kgs. All had eGFRs >80ml/min on day 0 and none had serum bilirubin, alkaline phosphatase or alamase aminotransferase enzyme levels of more than three times normal. Three of the five had co-morbidities – that is, cardiac, neurological or neoplastic disease. All HCQ serum concentrations were ≤0.005mg/L either before or on the first day of dosing. Serum concentrations on subsequent days after starting treatment are shown on Figure 1.

**FIGURE 1.**
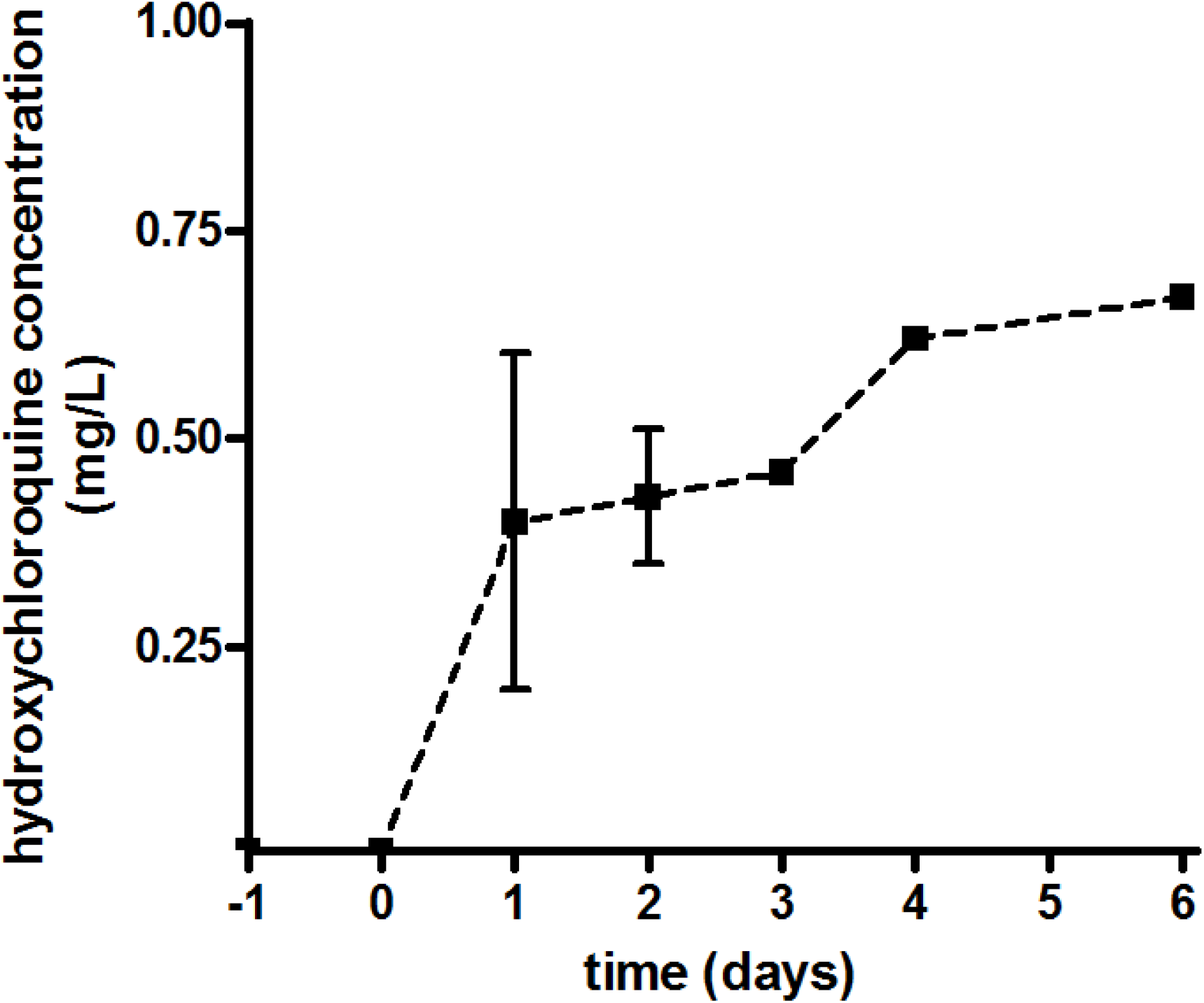
Serum concentrations of hydroxyl chloroquine.

Concentrations on day 1, 2, 3 and 4 were 0.4 ± 0.35mg/L (n=3); 0.43 ± 0.14mg/L (n=3), 0.46 mg/L (n=2), and 0.62mg/L (n=2) respectively.

The timing of sampling after oral dose was variable – 2.9 ± 3.5 (0.12-8.5 hours), though three quarters were taken within 4 hours of the dose being administered.

## Discussion

Data on the pharmacokinetics of HCQ has been accumulated in patients with conditions other than SARS-CoV-2 infection. The European SmPC suggest that after a single 400mg dose, the peak serum concentation was 0.105mg/L at 1.8 hr post dose (SmPC, 2017). Pharmacokinetic and modelling data exists for SARS-CoV-2 infected patients. Yoa et al, 2020 simulated several potential doses of HCQ suggesting that in Chinese subjects (usually smaller and lighter than European or North American patients) that a dose of 400mg BD and 200mg BD may be optimal. Interestingly, mean serum HCQ concentrations were not predicted to be >1mg/L until 4-5 days therapy with this regimen. Downes et al, 2020 simulated doses more akin to those used in the RECOVERY trial – that is, 1600mg on day 1 and 400mg/day thereafter. The predicted Cmax (IQR) was 2.7mg/L (2.2-3.3) and AUC 109.9mg/L.h. Perinel et al, 2020 performed a study of HCQ serum concentrations in French patients in critical care receiving 200mg TDS; most were mechanically ventilated. It took 2-7 days for serum concentration to reach 1mg/L. They also suggested a reference range of >1mg/L and <2mg/L but only 8 out of 13 patients reached these concentrations. HCQ dosing in the RECOVERY trial is higher than those in these studies and, despite this, all our patients, none of whom were receiving critical care, had HCQ concentration of <1mg/L. It may be expected that patients not receiving critical care may have better absorption than those in ICU.

There are a number of limitations of our study – most obviously, the small number of patients and samples available. However, our data indicates that HCQ concentrations are lower than might be expected, even with a high dose regimen.

Given that HCQ is likely to have marginal pharmacodynamics in SARS-CoV-2 infection, this data may in part help explain the lack of benefit in randomised clinical trials.

## Data Availability

Data available is accessible by request to the lead author

## Transparency declarations

Bristol Centre for Antimicrobial Research & Evaluation (BCARE) receives grant funding for research on antimicrobials from Paratek, Wockhardt, InfectoPharm, Venatorx, Merck, AiCURIS, NosoPharm, Evotec, IMI JU, COMBACTE MAGNET and GNA NOW, MRC(UK) and NIHR(UK).

